# Impact of Educational Resources for Primary Care OSA Management on Referral Patterns

**DOI:** 10.64898/2026.01.21.26344482

**Authors:** Shaelynn Zouboules, Willis H. Tsai, Oliver David, Ada Ip-Buting, Marcus Povitz, Reg Gerlitz, Gabriel E. Fabreau, Jaana Woiceshyn, W. Ward Flemons, Maoliosa Donald, Kerry McBrien, Sachin R. Pendharkar

## Abstract

**Background/Objective:** Barriers in access to care have prompted development of innovative care delivery models for patients with obstructive sleep apnea (OSA). One such innovation is management of uncomplicated OSA by primary care providers (PCPs), which reserves specialist capacity for more complex patients. We developed a clinical guideline and subsequently, an online clinical pathway to support PCPs in OSA management. We aimed to evaluate the impact of these initiatives on PCP behaviour by assessing changes in the complexity of newly referred patients.

**Methods:** We compared data for new OSA referrals to the Foothills Medical Centre Sleep Centre in Calgary, Canada, received during three distinct referral periods: prior to the clinical guideline (November 2016 – March 2017; Period 1), after implementation of the clinical guideline (April 2017 – November 2018; Period 2), and after publication of the online primary care pathway (December 2018 – November 2019; Period 3). The primary outcome was OSA severity as defined by the oxygen desaturation index (ODI) on home sleep apnea testing performed at the time of referral. Secondary outcomes included severity of daytime sleepiness and proportion of patients with severe OSA or severe daytime sleepiness at the time of referral. Multivariable linear and logistic regression models were constructed to quantify the associations between study outcomes and Period after adjustment for baseline covariates.

**Results:** Among the 2489 patients with sleep study data available, patients referred in Period 3 had more severe OSA (ODI, mean nocturnal oxygen saturation, proportion with severe OSA) compared to patients referred in earlier Periods. Severity of sleepiness did not increase across Periods. In multivariable analysis adjusting for demographics and comorbidities, ODI was significantly associated with Period (regression coefficient 1.70 (0.39, 3), p = 0.011) but severe OSA was not (odds ratio 1.17 (0.99, 1.39), p = 0.069).

**Conclusions:** OSA severity increased following implementation of educational resources to support primary care OSA management. These findings suggest that with appropriate supports, PCPs may be more comfortable managing less severe OSA patients independently and only refer more severe cases for specialist consultation.

## INTRODUCTION

Obstructive sleep apnea (OSA) is a chronic sleep disorder characterized by recurrent complete or partial upper airway obstruction. Globally, over 900 million adults aged 30-69 are estimated to have OSA. (1–3) Despite its high prevalence in population-based studies, OSA is underdiagnosed in North America, with only 6.4% of Canadians reporting a formal diagnosis despite 30% being at risk. (4–6) There are numerous consequences of untreated OSA including: increased cardiometabolic risk, motor vehicle collisions, occupational injuries and poor quality of life. (7) Furthermore, individuals with OSA use more healthcare resources and are at higher risk of hospitalization compared to the general population. (7–9) Treatment of OSA both improves health outcomes and is cost-effective. (7,10)

Barriers to accessing timely sleep specialist care for OSA have prompted interest in the development of innovative care delivery models. One such model is a community-based care model with primary care providers (PCPs) at the centre, which can improve access for most patients with less complicated OSA and reserve specialty care for more complex patients.(11) Several studies have demonstrated that, in a research setting, primary care management of uncomplicated OSA (i.e., without other sleep disorders or significant comorbidities) can achieve similar outcomes as specialist care. (12–16) However, enthusiasm for primary care OSA management must be balanced with evidence of knowledge gaps and poor confidence about OSA among PCPs (17–21). In prior consultation with PCPs and patients, we identified a need for educational resources to support PCPs to manage OSA.(22)

Prompted by long wait times for sleep specialist assessment, clinicians at the Foothills Medical Centre (FMC) Sleep Centre in Calgary, Alberta developed a clinical guideline entitled “Obstructive Sleep Apnea - Guidelines for Diagnosis and Treatment,” which was disseminated to all referring physicians starting in March 2017. We subsequently engaged primary care leaders in Calgary in the co-development of an online primary care clinical pathway (“Primary Care Pathway: Uncomplicated Obstructive Sleep Apnea”) to replace the OSA clinical guideline. The clinical pathway was published in December 2018 on Specialist LINK, a website that houses clinical pathways for a variety of chronic conditions and facilitates real-time telephone access to specialists in Calgary. Specialist LINK is frequently used by the 1700 urban and rural PCPs in Calgary and area (23).

These initiatives were intended to support PCPs to manage uncomplicated OSA but their influence on referring physician behaviour have not been evaluated. Therefore, the primary aim of this project was to determine the impact of the clinical guideline and pathway on the complexity of patient referrals to the FMC Sleep Centre.

## METHODS

### Study Design

This retrospective observational study was conducted at the FMC Sleep Centre in Calgary, Canada. We evaluated sociodemographic and clinical characteristics for new referrals received during three distinct Periods: prior to the clinical guideline, after implementation of the clinical guideline, and after dissemination of the primary care pathway via Specialist LINK (Figure 1). The Conjoint Health Research Ethics Board (CHREB) at the University of Calgary approved this study (ID # REB21-0433).

**Figure 1.**
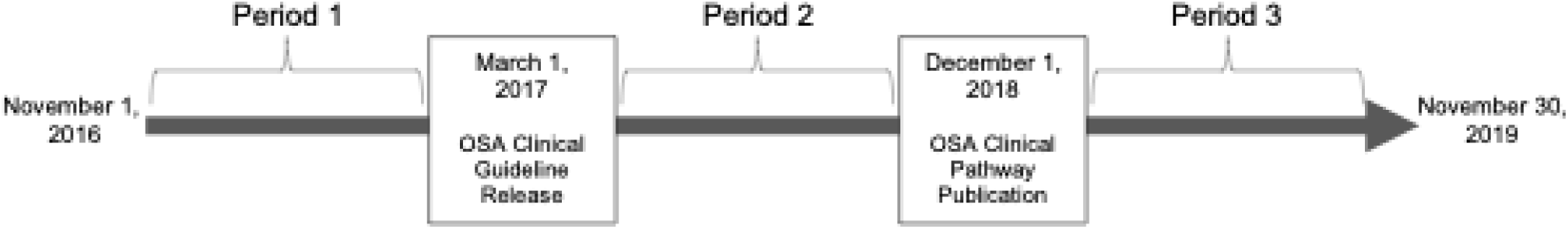
Schematic of study Periods. OSA – obstructive sleep apnea

### Study Setting

In Calgary, PCPs may refer patients with OSA to hospital-based or community sleep specialists, or manage them independently using community-based respiratory homecare companies who perform home sleep apnea testing (HSAT) and sell continuous positive airway pressure (CPAP) equipment. Polysomnography is available only in specialized sleep laboratories by specialist referral. Treatment options are typically funded privately and include CPAP therapy that is provided by respiratory homecare companies, mandibular advancement devices provided by dentists and selected upper airway surgical procedures performed by head and neck surgeons.

The FMC Sleep Centre is a tertiary academic sleep centre and the only public sleep centre in Calgary. At the FMC Sleep Centre, OSA care is provided by eight sleep physicians with support from five sleep-trained respiratory therapists. Approximately 2500 referrals are received each year from the Calgary Zone and Southern Alberta (catchment population of approximately 2 million), of which over 50% are for OSA (24). Newly referred patients undergo HSAT and complete a triage questionnaire to establish urgency and the most appropriate provider that should assess the patient.

Prior to the implementation of the access initiatives, all referrals to the FMC Sleep Centre were accepted and prioritized for sleep specialist assessment. After March 1, 2017, referrals for uncomplicated OSA were closed and returned to the PCP accompanied by the HSAT result and the clinical guideline. As of December 1, 2018, referrals for uncomplicated OSA continued to be closed but the clinical guideline was replaced by a link to the clinical pathway on the Specialist LINK website. After receiving the returned referral, the PCP could manage the patient independently, refer the patient to a community-based sleep specialist, or re-refer to the FMC Sleep Centre.

### Study Cohort and Data

We extracted referral and clinical data for all patients who were referred for suspected OSA between November 2016 to November 2019. Data were obtained from the FMC Sleep Centre electronic health record (Sleep Information System, SIS). This system contains all clinical, scheduling and operational data for the FMC Sleep Centre from November 2016 onwards.

Exclusion criteria included: referrals for sleep disorders besides OSA, referrals that were not triaged as having sleep-disordered breathing, provincial health number from outside of Alberta, or referrals with insufficient information to determine if OSA was the reason for referral. Since SIS was implemented in November 2016, we noted that the referral date field was populated by first visit date for some existing patients who had not been newly referred. To reconcile this issue, we validated the referral date using other data fields (e.g., visit history) to ensure patients were newly referred.

Variables of interest included: patient characteristics (age, sex, body mass index, neck circumference), baseline sleepiness using the Epworth Sleepiness Scale (ESS), and HSAT results (oxygen desaturation index [ODI], mean nocturnal oxygen saturation [SpO_2_]). Sleep testing results were only available for patients who had HSAT completed through the FMC Sleep Centre. Important clinical comorbidities such as hypertension, diabetes, asthma, COPD, depression, dementia, chronic kidney disease, cardiovascular disease, stroke and chronic pain were determined using ICD-9 CM/ICD-10 administrative health data based on validated algorithms in the province of Alberta (25).

### Analysis

We defined three time periods for analysis of the impact of each initiative (Figure 1). The start of Period 1 coincided with the implementation of SIS, which enabled us to obtain standardized referral data. Period 2 encompassed the time that the clinical guideline accompanied closed referrals and Period 3 included 12 months after the clinical pathway was published on Specialist LINK.

The primary outcome was severity of OSA, assessed using ODI on triage HSAT that was performed shortly after referral to the FMC Sleep Centre. Secondary outcomes included mean nocturnal SpO_2_, ESS score, and proportion of patients with severe OSA (ODI ≥ 30/hour) or severe sleepiness (ESS ≥16).

We used descriptive statistics to describe patient characteristics in each Period. Analysis of variance (ANOVA) and chi squared tests were used to compare unadjusted study outcomes across Periods. We then constructed multivariable linear and logistic regression models to investigate the association of study outcomes and intervention Period after adjusting for age, sex, body mass index (BMI), neck circumference, comorbidities and median neighbourhood income. As individual sociodemographic data were not available in our data sources, postal codes were linked to Statistics Canada’s Postal Code Conversion File to obtain aggregate neighborhood data (26).

## RESULTS

There were 3622 referrals received during the entire study. Mean (standard deviation, SD) age at referral was 55.6 (14.9) years and 59% were male. Mean (SD) BMI was 32.7 (8.25) kg/m2. There was no difference between age, sex, or comorbidity across Periods. Body mass index increased from Period 1 to Period 3 (Table 1).

**Table 1:** Patient characteristics.

| Variable | Period 1<br>(n = 483) | Period 2<br>(n = 2007) | Period 3<br>(n=1132) | p value* |
| --- | --- | --- | --- | --- |
| Age, years | 54.5 (14.5) | 55.4 (14.7) | 56.2 (15.4) | 0.18 |
| Male sex, n (%) | 294 (61) | 1190 (59) | 669 (59) | 0.79 |
| BMI, kg/m<sup>2</sup>* | 31.8 (7.4) | 32.4 (8) | 33.7 (9) | < 0.001 |
| Neck circumference, cm† | 40.8 (5.4) | 41 (5) | 41.6 (5.2) | 0.28 |
| Median neighbourhood income, CAD‡ | 44847 (11567) | 44349 (11224) | 43981 (10802) | 0.17 |
| Comorbidities, n (%) |  |  |  |  |
| Asthma | 41 (8.5) | 154 (7.7) | 89 (7.9) | 0.84 |
| Atrial fibrillation | 31 (6.4) | 134 (6.7) | 75 (6.6) | 0.98 |
| Chronic Pain | 62 (12.8) | 289 (14.4) | 152 (13.4) | 0.58 |
| COPD | 57 (11.8) | 205 (10.2) | 114 (10.1) | 0.54 |
| Dementia | 6 (1.2) | 32 (1.6) | 17 (1.5) | 0.85 |
| Depression | 103 (21.3) | 418 (20.8) | 227 (20) | 0.81 |
| Diabetes mellitus | 106 (21.9) | 440 (21.9) | 263 (23.2) | 0.68 |
| Heart Failure | 34 (7.0) | 154 (7.7) | 95 (8.4) | 0.61 |
| Hypertension | 183 (37.9) | 849 (42.3) | 482 (42.6) | 0.17 |
| Myocardial infarction | 8 (1.7) | 37 (1.8) | 26 (2.3) | 0.59 |
| Kidney Disease | 38 (7.9) | 176 (8.8) | 108 (9.5) | 0.54 |
| Peripheral Vascular Disease | 10 (2.1) | 29 (1.4) | 16 (1.4) | 0.57 |
| Stroke | 10 (2.1) | 48 (2.4) | 19 (1.7) | 0.41 |

1. Data are presented as mean (standard deviation) unless otherwise specified.
2. BMI – body mass index; CAD – Canadian dollars; COPD – chronic obstructive pulmonary disease. \*n = 2269 patients; †n = 2224 patients; ‡n=3471 patients.

Results of sleep testing and OSA symptoms were available for 2489 and 2232 patients, respectively (Table 2 and Figure 2). In Period 2, we did not identify a change in any study outcomes compared to Period 1. However, after launch of the online OSA pathway (Period 3), measures of OSA severity (ODI, mean nocturnal SpO2, proportion with severe OSA) all indicated more severe disease compared to each of the prior two Periods. There were no statistically significant differences in ESS or proportion of patients with severe sleepiness across Periods. In multivariable analysis including demographic characteristics and comorbidities, ODI was significantly associated with Period (regression coefficient 1.70 (0.39,3), p = 0.011) but severe OSA was not (odds ratio 1.17 (0.99, 1.39), p = 0.069).

**Figure 2.**
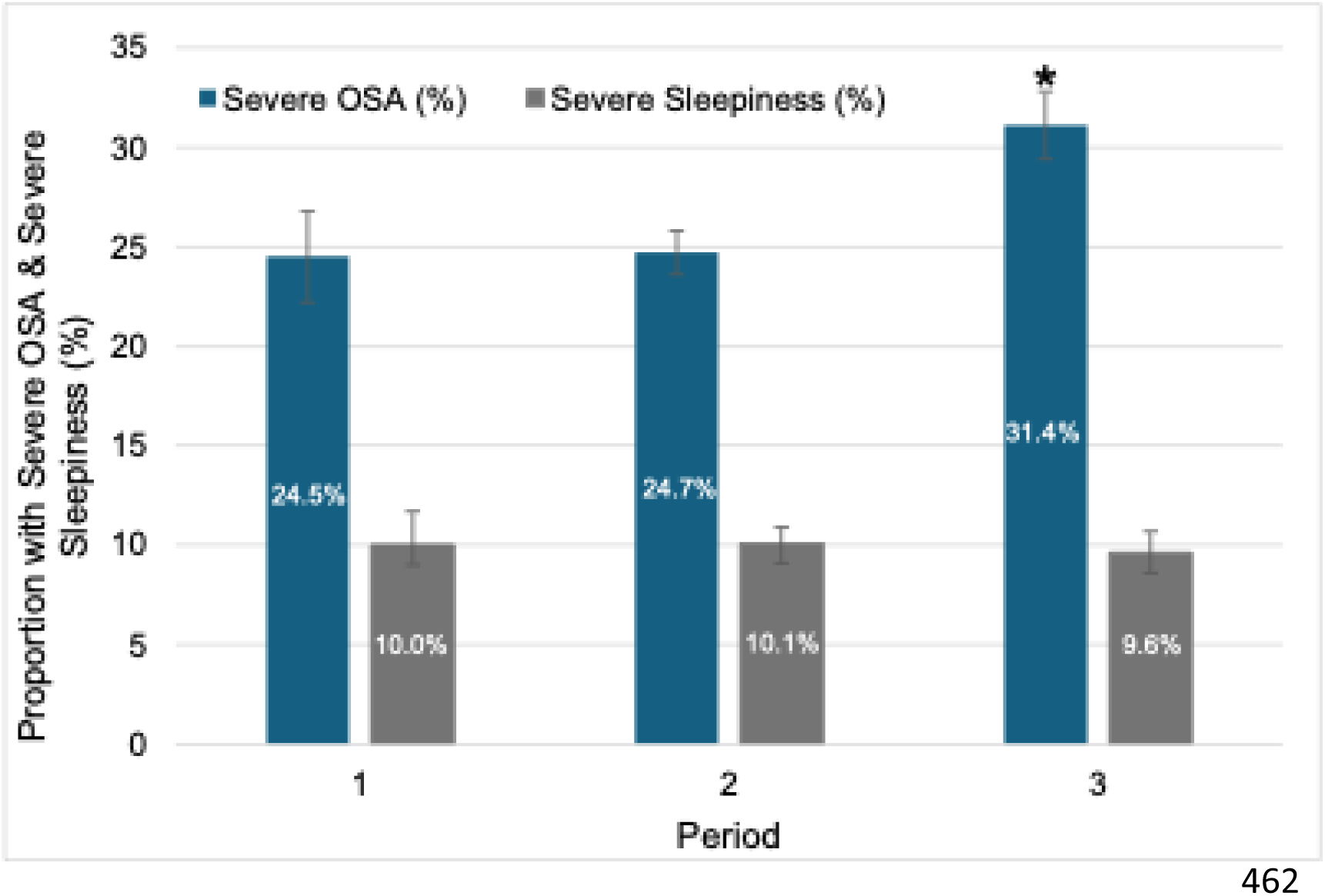
Proportion of patients referred in each Period with severe obstructive sleep apnea (OSA) or severe sleepiness. *Statistically significant difference (P<0.01) across Periods.

**Table 2:** Unadjusted study outcomes.

| Variable | Period 1<br>(n = 327) | Period 2<br>(n = 1403) | Period 3<br>(n = 759) | p value |
| --- | --- | --- | --- | --- |
| **ODI, events/h** | 20.5 (21.6) | 22.1 (23.9) | 25.9 (25.4)<sup>†‡</sup> | <0.001 |
| **Severe OSA (ODI ≥ 30), n (%)** | 80 (24.5) | 347 (24.7) | 238 (31.4) | <0.01 |
| **ESS score\*** | 7.9 (5.2) | 8.1 (5.1) | 7.8 (5.2) | 0.538 |
| **Severe Sleepiness (ESS ≥16), n (%)\*** | 31 (10) | 127 (10) | 62 (9.6) | 0.090 |
| **Mean nocturnal SpO<sub>2</sub>, %** | 90.5 (5.2) | 90 (3.9) | 89.4 (4.3) | <0.01 |

1. Data are presented as mean (standard deviation) unless otherwise indicated.
2. ODI - oxygen desaturation index; OSA - obstructive sleep apnea; ESS - Epworth Sleepiness Scale; SpO<sub>2</sub> - oxygen saturation. \*n = 2232.
3. Overall P-values: continuous variables (ODI, Mean nocturnal SpO<sub>2</sub>, ESS) tested using regression/ANOVA across periods; categorical variables (Severe OSA, ESS ≥16) tested using chi-square.
4. Symbols for pairwise comparisons: <sup>†</sup>P<0.01 vs Period 1; <sup>‡</sup>P<0.01 vs Period 2 (ODI only, as pairwise comparisons performed).

## DISCUSSION

The objective of this study was to evaluate the impact of two sequentially implemented educational resources that aimed to support primary care management of uncomplicated OSA in Calgary: a clinical guideline sent to referring physicians and an online clinical pathway that was co-designed with end users. Overall, we found that the dissemination of these resources was associated with a significant increase in OSA severity based on HSAT-derived metrics, with a possible specific benefit of the online pathway that was launched in Period 3. Symptoms did not change significantly. To our knowledge, this is the first study to demonstrate the potential system benefits of educational resources such as a clinical pathway for OSA.

In many jurisdictions, assessment and management of OSA is performed by sleep specialists, even though the prevalence of OSA is comparable to other chronic conditions typically managed in primary care settings. The high prevalence of OSA, combined with the limited supply of sleep specialists, has resulted in unacceptably long wait times for OSA diagnosis and treatment, (27–29) which may lead to undertreatment, poor treatment adherence and frustration with care. (11,17,30–32) Moreover, a model that relies on sleep specialists poses additional barriers to individuals who already have difficulty accessing specialist care, such as those living in rural communities or who are socially vulnerable. (17,33–38) This traditional, specialist-centered model is both unsustainable and inequitable, and has prompted calls to improve the design and implementation of non-specialist models in OSA care. (11,31,32) The management of uncomplicated OSA in a primary care setting has been demonstrated to be non-inferior to specialist management and cost-saving. (12–15) As such, primary care management has been suggested as a means to improve access to care for patients with OSA. (16,17) However, barriers to implementation in clinical practice include knowledge gaps and poor confidence about OSA management among PCPs. (17,21) Educational tools such as a clinical pathway provide an opportunity to overcome these barriers. Prior randomized trials of OSA management by PCPs have included OSA-specific education sessions, but our evaluation suggests that co-developed educational resources can support PCPs to manage uncomplicated OSA outside of research settings.

Our findings build on results from studies evaluating other disease-specific pathways and other tools to support collaborative care between PCPs and specialists. Recently, pathways from the Calgary Specialist LINK website have been evaluated, demonstrating high awareness and uptake as well as positive impacts on patient outcomes. (39,40) Other strategies include assessment tools embedded in electronic medical records, (41) or communication platforms such as the Canadian Foundation for Healthcare Improvement’s “Connected Medicine collaborative,” which comprises both an asynchronous online messaging platform and a synchronous telephone service. (42) These strategies have strong potential to reduce access barriers and improve system outcomes such as unnecessary specialist referral or emergency department visits. (42)

We defined each Period based on the timing of the associated intervention (clinical guideline and online pathway). Our results demonstrate a statistically significant increase in OSA severity during Period 3 compared to Periods 1 and 2. It is possible that these findings reflect greater awareness and use of the online clinical pathway due to co-development with PCPs and publication on an easily accessible and widely used website (Specialist LINK). However, it is also possible that results in Period 3 reflect a delayed effect of the clinical guideline since the resources were provided to PCPs at the time that patients were referred rather than as discrete interventions at the beginning of a Period. Given the time required for behaviour change interventions to influence clinical practice,(43) our results may represent the collective impact of sequential interventions to support primary care OSA management.

Strengths of our study include the large sample size and that referral patterns were assessed over years, with the study design incorporating distinct time periods to attempt to individually assess each initiative. We must also acknowledge limitations, most notably that we did not measure fidelity to the interventions (pathway use), so we cannot directly link the increase in OSA severity to uptake of the resources. It is possible that the demonstrated associations are coincidental and related to other drivers of change in referral patterns such as an increase in the number of community homecare companies, an aging population, or other secular trends. The uncontrolled study design precludes determination of the contributions of these and other factors. Second, the FMC Sleep Centre began returning referrals (accompanied by the clinical guideline) in Period 2; thus, it is possible that independent management of OSA was prompted by this action rather than either the clinical guideline or online pathway. Future studies, including qualitative studies of PCPs, could explore the drivers of PCP referral behaviours for OSA in more detail. Finally, although there was no between group difference in postal code derived variables between Periods, the use of aggregate data in regression models may lead to incorrect adjustment of associations for individual patients (ecological fallacy).

## CONCLUSION

In this retrospective observational study, we demonstrated an association between sequential implementation of a clinical guideline and online clinical pathway for OSA management and greater severity of OSA at the time of specialist referral. Given increasing demands on PCPs in a fragmented and resource constrained health system, these findings suggest that such educational interventions may provide needed supports for primary care management of OSA and improved access to OSA care.

## Data Availability

The data underlying this study cannot be shared due to research ethics board requirements which protect the confidentiality of the research participants.

